# Post head and neck cancer radiation therapy dynamic contrast enhanced magnetic resonance imaging association with high mandibular radiation dose

**DOI:** 10.1101/2025.07.25.25332216

**Authors:** Brandon Reber, Renjie He, Moamen R Abdelaal, Abdallah Mohamed, Sam Mulder, Laia Humbert Vidan, Clifton D. Fuller, Stephen Y. Lai, Kristy Brock

**Affiliations:** Department of Imaging Physics, The University of Texas MD Anderson Cancer Center, Houston, Texas, US0041; Department of Radiation Oncology, Mayo Clinic, Rochester, Minnesota, USA; Department of Radiation Oncology, The University of Texas MD Anderson Cancer Center, Houston, Texas, USA; The University of Rochester, Rochester, New York, USA; Department of Head and Neck Surgery, The University of Texas MD Anderson Cancer Center, Houston, Texas, USA; 6 Department of Radiation Physics, The University of Texas MD Anderson Cancer Center, Houston, Texas, USA

**Keywords:** Head and neck cancer, dynamic contrast-enhanced MRI, mandible, radiation

## Abstract

**Background:** Dynamic contrast enhanced magnetic resonance imaging (DCE-MRI) is a functional imaging modality that can quantify tissue permeability and blood flow. Due to vasculature changes resulting from radiation therapy (RT), DCE-MRI quantitative parameters should be significantly different in regions receiving a high radiation dose compared to regions receiving a low radiation dose. This work sought to determine if a significant difference exists in post head and neck cancer (HNC)-RT DCE-MRI quantitative parameters K^trans^ and v_e_ between regions of the mandible receiving a high radiation dose and regions of the mandible receiving a low radiation dose.

**Methods:** DCE-MRI was acquired from HNC subjects post-RT. The DCE-MRI quantitative parameters K^trans^ and v_e_ were obtained through Tofts model fitting. Four mandible sections (left ramus, left body, right ramus, and right body) were delineated on subject mandible contours. Two Kruskal-Wallis tests comparing the mean K^trans^ and v_e_ in low dose (≤ 60 Gy) areas of the four mandible regions were computed. Next, two Wilcoxon signed-rank tests were used to determine if the means of K^trans^ and v_e_ between high dose (> 60 Gy) and low dose (≤ 60 Gy) mandible regions were significantly different. To account for multiple statistical tests, a Bonferroni corrected significance level for all statistical tests was used 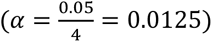.

**Results:** 48 HNC subjects were included in the analysis. The Kruskal-Wallis tests showed no inherent significant difference in K^trans^ means between mandible regions 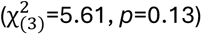 and no inherent significant difference in v_e_ means between mandible regions 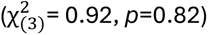. No significant difference was found between high and low dose K^trans^ mandible means (*W*=392, *p*=0.044). A significant difference was found between high and low dose v_e_ mandible means (*W*=214, *p*=0.00013).

**Conclusions:** No inherent difference in DCE-MRI quantitative parameters was observed within subject mandibles, but a significant difference was observed between v_e_ means of high and low radiation dose mandible regions. These results provide evidence of the utility of DCE-MRI to monitor mandible vasculature changes resulting from head and neck cancer radiation therapy. Monitoring post HNC-RT mandible vasculature changes is important to initiate earlier toxicity management and ultimately improve HNC survivor quality of life.

**Simple Summary:** Dynamic contrast-enhanced magnetic resonance imaging (DCE-MRI) can detect relative differences in anatomical blood perfusion and vessel permeability. Differences in vascularity should occur between mandible regions receiving a large radiation dose and regions receiving a low radiation dose during head and neck cancer radiation therapy. In this study, we determined if DCE-MRI can be used to detect vasculature differences between mandible regions irradiated with high and low amounts of radiation. The results indicate that one of the DCE-MRI parameters was significantly different between irradiated mandible regions receiving high and low radiation dose. This parameter may be used as an early marker for mandible radiation damage from head and neck radiation therapy.

## INTRODUCTION

In 2020 and 2021, about 660,000 patients were diagnosed with head and neck cancer (HNC), and 325,000 HNC-related deaths occurred worldwide[1][2]. HNCs form in the mucosal surfaces of the pharynx, larynx, oral cavity, paranasal sinuses, and salivary glands[3], with most being squamous cell carcinomas[4]. Risk factors for HNC vary depending on the disease subsite, but they generally include tobacco use, alcohol consumption, human papillomavirus status, and overall oral health[5]. Owing to an increase in HNC associated with human papillomavirus, incidence rates for HNC are expected to increase 30% annually from 2018 to 2030[2].

The most common treatment method for head and neck squamous cell carcinoma is a combination of surgery, chemotherapy, and radiation therapy (RT)[6][7]. The delivered radiation dose depends on several factors, but it typically ranges from 50 Gy to 70 Gy[7]. Several toxic effects, such as xerostomia and dysphagia, can occur during or after RT[8]. Osteoradionecrosis (ORN) is a toxic effect that can result from radiation exposure across the volume of the mandible, especially in the treatment of oral cavity and oropharyngeal cancers[9][10]. Given the cumulative lifetime risk for ORN and the resulting impact on patient function and quality-of-life, identifying patients at risk for ORN or already suffering from early-stages of ORN is critical[11][12].

Dynamic contrast-enhanced (DCE)-MRI is a functional imaging modality that can measure blood perfusion, vascularity, and permeability in regions of interest[13]. DCE-MRI involves the injection of a contrast agent that alters the measured MRI signal in regions adjacent to the contrast agent[14]. These changes in signal intensity are ultimately related to differing blood perfusion and tissue permeability within the imaged regions[14]. Several pharmacokinetic models are available to determine the physiological relationship to measured signal intensities. One of the most commonly used pharmacokinetic models is the Tofts model[15]. This model defines several quantitative parameters, including K^trans^, the transfer constant from plasma to the extravascular extracellular space (EES), and v_e_, the fractional volume of the EES[15]. According to the Tofts model, differences in these parameters between regions indicate relative differences in tissue permeability and blood perfusion. Because RT-damaged tissues can have changes in blood perfusion and tissue permeability[16][17], DCE-MRI quantitative parameters related to perfusion and tissue permeability should differ among radiation-damaged and non–radiation-damaged tissues.

Previously, researchers have used DCE-MRI for HNC imaging, such as for segmenting HNCs [14], HNC tumor staging and grading[18], histopathology correlation[19], and treatment response monitoring[20]. Some previous studies have looked at using DCE-MRI to characterize the vasculature and perfusion changes within the mandible resulting from HNC-RT[21][22][23][24][25]. In one study, investigators examined the changes in DCE-MRI parameters before and after treatment in the mandibles of rabbits and found that DCE-MRI may be able to model maxillofacial wound healing[25]. In another study, researchers compared K^trans^ and v_e_ in regions of the same mandible that did and did not have osteoradionecrosis and[24] found that the K^trans^ and v_e_ were significantly different between ORN affected and ORN free regions[24]. Another study demonstrated significant voxel-wise differences in the K^trans^ and v_e_ using DCE-MRI before and after RT[22]. However, to date, no study has looked at differences in DCE-MRI parameters in different mandibular regions irradiated with high and low radiation dose not necessarily related to observable ORN.

In this work, we sought to determine if DCE-MRI can detect changes in the permeability and blood perfusion in the mandible as a result of HNC-RT. To this end, we analyzed the post-RT means of K^trans^ and v_e_ in high-dose mandible regions (>60 Gy) and those in low-dose mandible regions (≤60 Gy). Owing to tissue damage resulting from RT, K^trans^ and v_e_ should differ significantly between the regions because of differences in blood perfusion and tissue permeability.

## METHODS

### Overview

The overall research methodology was split into 3 parts. First, DCE-MRI quantitative parameters K^trans^ and v_e_ were collected, curated, and registered. Next, the parameters for the low-dose mandible regions were analyzed for inherent significant differences in K^trans^ and v_e_ within the mandible. Finally, the means of K^trans^ and v_e_ in the high-dose mandible regions were compared to parameter means in low-dose mandible regions. The second and third methodology components are summarized in Figure 1.

**FIGURE 1.**
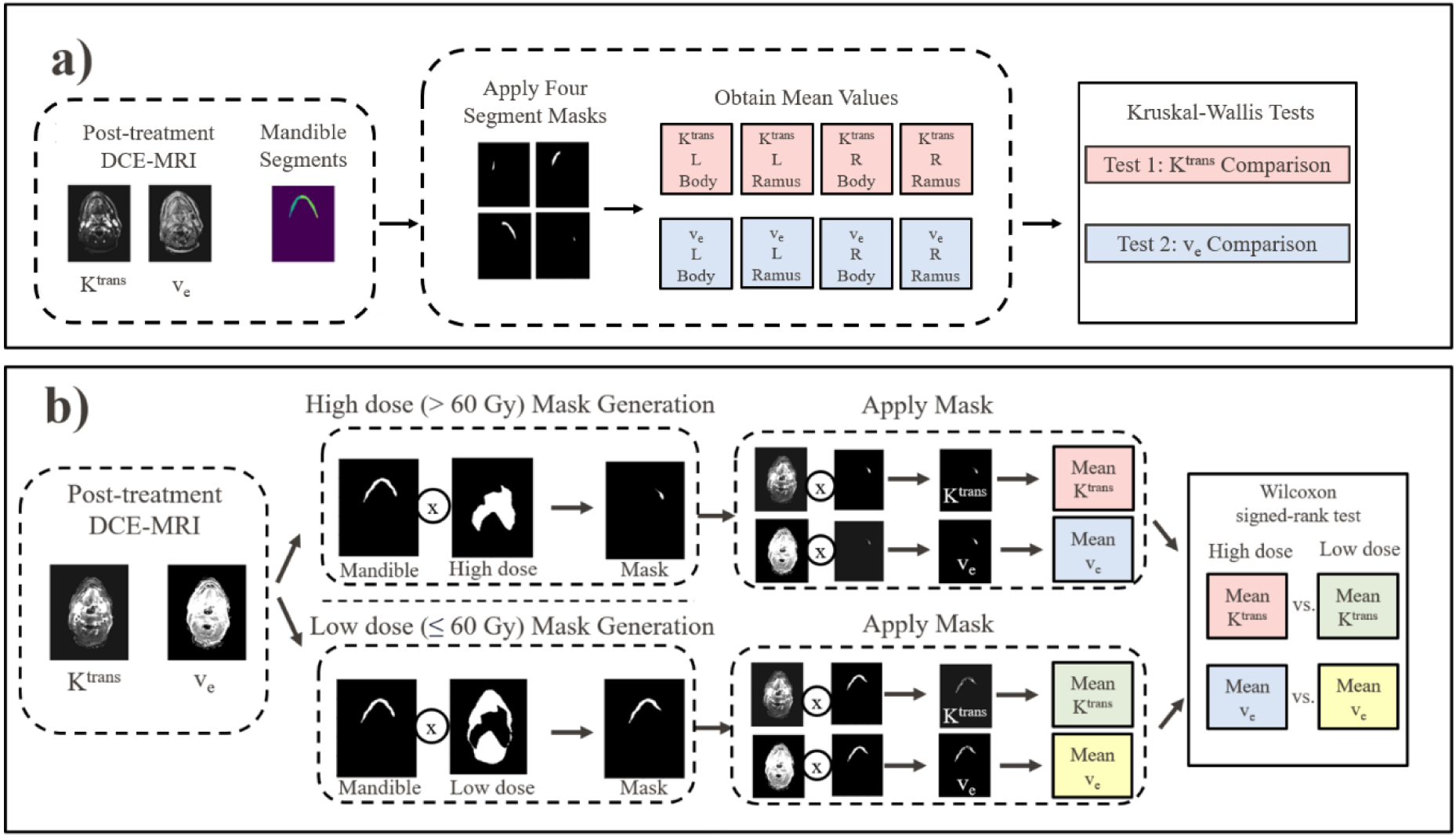
Overview of the research methodology. a) shows the comparison process between the DCE-MRI of the four mandible regions. This was completed to determine if an inherent difference in DCE-MRI exists between the four mandible regions unrelated to changes associated with tissue damage from radiation therapy. b) illustrates the DCE-MRI comparison between high and low dose regions of the mandible. This process determines if the DCE-MRI can capture changes in mandibular K^trans^ and v_e_ associated with radiation therapy.

### Patients cohort

Patients were included from an ongoing clinical trial at The University of Texas MD Anderson Cancer (clinicaltrials.gov ID: NCT03145077). The trial enrollment eligibility criteria were patients with age older than 18 years, curative RT for HNC, ability to undergo MRI, and an Eastern Cooperative Oncology Group (ECOG) performance status score of 0-2. Patients received prescription doses ranging from 60 Gy to 70 Gy delivered in 30 to 35 fractions. All patients underwent follow-up DCE-MRI at least 1 month after RT completion. Patients were excluded if they received treatment with multiple modalities, such as a combination of intensity-modulated RT and intensity-modulated proton therapy.

### Data

Patient RT dose maps, gross tumor volume (GTV) contours, and treatment planning CT scans were acquired from a clinical database in RayStation 11B (RaySearch Laboratories, Stockholm, Sweden). T2-weighted images were acquired using a Siemens Aera 1.5T MRI scanner (Siemens Heathineers, Erlangen, Germany; TE/TR = 80/4800 ms, matrix size = 512 × 512, slice thickness = 2 mm, voxel spacing = 0.5 mm × 0.5 mm, and 1 average) during the post-treatment DCE-MRI (TE/TR = 1.07/5 ms, matrix size = 256 × 208, slice thickness = 4 mm, voxel spacing = 1 mm × 1 mm, 1 average). The contrast agent gadobutrol (Gadovist; Bayer Healthcare, Leverkusen, Germany) was injected with a power injector (Spectris MR Injector; MedRad, Pittsburgh, PA) at a dose of 0.1 mmol/kg body weight at 3 ml/s. In combination with the contrast agent, saline was administered at 3 ml/s at the same quantity as that of the contrast agent. The pharmacokinetic modeling procedure used to fit the DCE-MRI was described previously[26]. Briefly, the Tofts model was used, which can be defined as follows[15]:

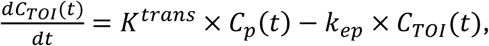

where 𝐶_𝑇𝑂𝐼_(𝑇) is the concentration of the contrast agent in the tissue of interest, 𝐾^𝑡𝑟𝑎𝑛𝑠^is the volume transfer constant of the contrast agent from the plasma into the EES, 𝐶_𝑝_(𝑡) is the contrast agent in the plasma, and 𝑘_𝑒𝑝_(𝑡) is the transfer constant of contrast agent from the plasma into the EES. 𝑘_𝑒𝑝_(𝑡) is related to the volume fraction of the EES through the following formula:

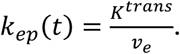

The arterial input function used in the pharmacokinetic model fitting was obtained via fitting a 7- parameter biexponential, bilinear arterial input model function[26]. The pharmacokinetic modeling was completed on a voxel-to-voxel basis.

### Registration

Rigid registration was performed to align each patient’s DCE-MRI and treatment dose maps. The DCE-MRI map was in the same frame of reference as that of the T2-weighted image, and the treatment dose map was in the same frame of reference as that of the treatment planning CT. The rigid registration was completed between patient’s T2-weighted and treatment planning CT images. The T2-weighted image was used as the fixed image, and the CT image was used as the moving image. The intensity-based rigid registration was completed using RayStation 11B. A mandible contour of the head and neck was generated on each patient’s treatment planning CT image using an atlas-based segmentation in RayStation 11B. SimpleITK (version 2.3.0) was then used to resample the CT image, dose map, GTV contour, and mandible contour to the spacing of the DCE-MRI parameters using linear interpolation. The resulting outputs were the treatment planning CT, mandible contour, GTV contour, and dose images that were registered and resampled to the K^trans^ and v_e_ images.

### Comparison of DCE-MRI in different mandible regions

Regions of interest in the mandible corresponding to the left ramus, left body, right body, and right ramus were delineated by a graduate student on each patient’s treatment planning CT map. Next, voxels were removed from the K^trans^ and v_e_ regions of the mandible that corresponded to either high-dose (>60 Gy) or GTV regions. These voxels were removed to test inherent DCE-MRI differences between mandible regions while limiting the potential effect of high dose or the GTV on DCE-MRI parameters. The mean values of voxels within each of the 4 regions of interest for each patient were then collected for the K^trans^ and v_e_ parameters separately. In all, 8 mean values were collected for each patient: K^trans^ left ramus, K^trans^ left body, K^trans^ right body, K^trans^ right ramus, v_e_ left ramus, v_e_ left body, v_e_ right body, and v_e_ right ramus (Figure 1a).

### High- and low-dose volume selection

After DCE-MRI comparisons between mandible regions, separate additional comparisons were completed between DCE-MRI high dose (> 60 Gy) and low-dose (≤60 Gy) mandible regions. The dose threshold of 60 Gy was chosen due to prior evidence of mandibular doses > 60 Gy being associated with an increased ORN risk[27]. Two binary dose masks were created using patient dose masks to select the high-dose and low-dose regions of the mandible. The GTV portions of the mandible were removed from the masks. The GTV was removed from DCE-MRI masks to limit the effects of DCE-MRI changes resulting from the GTV. The means of the high- and low-dose volumes were then computed for both the K^trans^ and v_e_ parameters separately. In all, 4 mean values were generated for each patient: high-dose K^trans^, low-dose K^trans^, high-dose v_e_, and low-dose v_e_ (Figure 1b).

### Statistical analysis

Two sets of statistical tests were completed. The first set of tests compared the low dose areas of the four mandible regions to determine if an inherent significant difference in DCE-MRI exists within the mandible. This set of tests comprised of two Kruskal-Wallis tests. One test compared the K^trans^ of the four mandible groups and the other test compared the v_e_ of the four mandible groups. Next, a second set of tests was completed to determine if a significant difference exists between the high and low dose DCE-MRI regions. This set of tests comprised two Wilcoxon-signed rank tests which compare per patient DCE-MRI differences. One test compared the high and low dose K^trans^ groups and one test compared the high and low dose v_e_ groups. A Bonferroni correction was applied to address multiple comparisons for all tests. The adjusted significance level was 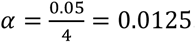. Therefore, statistical significance was determined when *P* < 0.0125.

## RESULTS

A total of 48 subjects were included in the study. A summary of the 48 patients’ demographics is presented in Table 1.

**TABLE 1.** Patient demographic characteristics (n=48).

| Characteristic | n (%) [IQR] |
| --- | --- |
| Median age, years | 64 [13] |
| Male sex | 43 (90%) |
| Current smoker | 1 (2%) |
| Former smoker | 28 (58%) |
| Median packs per year, n |  |
| Current smoker | 18 [0] |
| Current and former smokers | 15 [12.5] |
| Tumor site |  |
| Oral cavity | 6 (13%) |
| Oropharynx | 40 (83%) |
| Other* | 2 (4%) |

\*Other tumor sites were the hypopharynx, larynx, nasopharynx, and unknown.

Boxplots of the K^trans^ and v_e_ values for the left ramus, left body, right body, and right ramus are shown in Figure 2. We found no significant differences between the parameters and mandible regions using the two Kruskal-Wallis tests 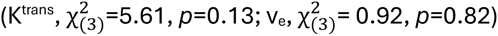.

**FIGURE 2.**
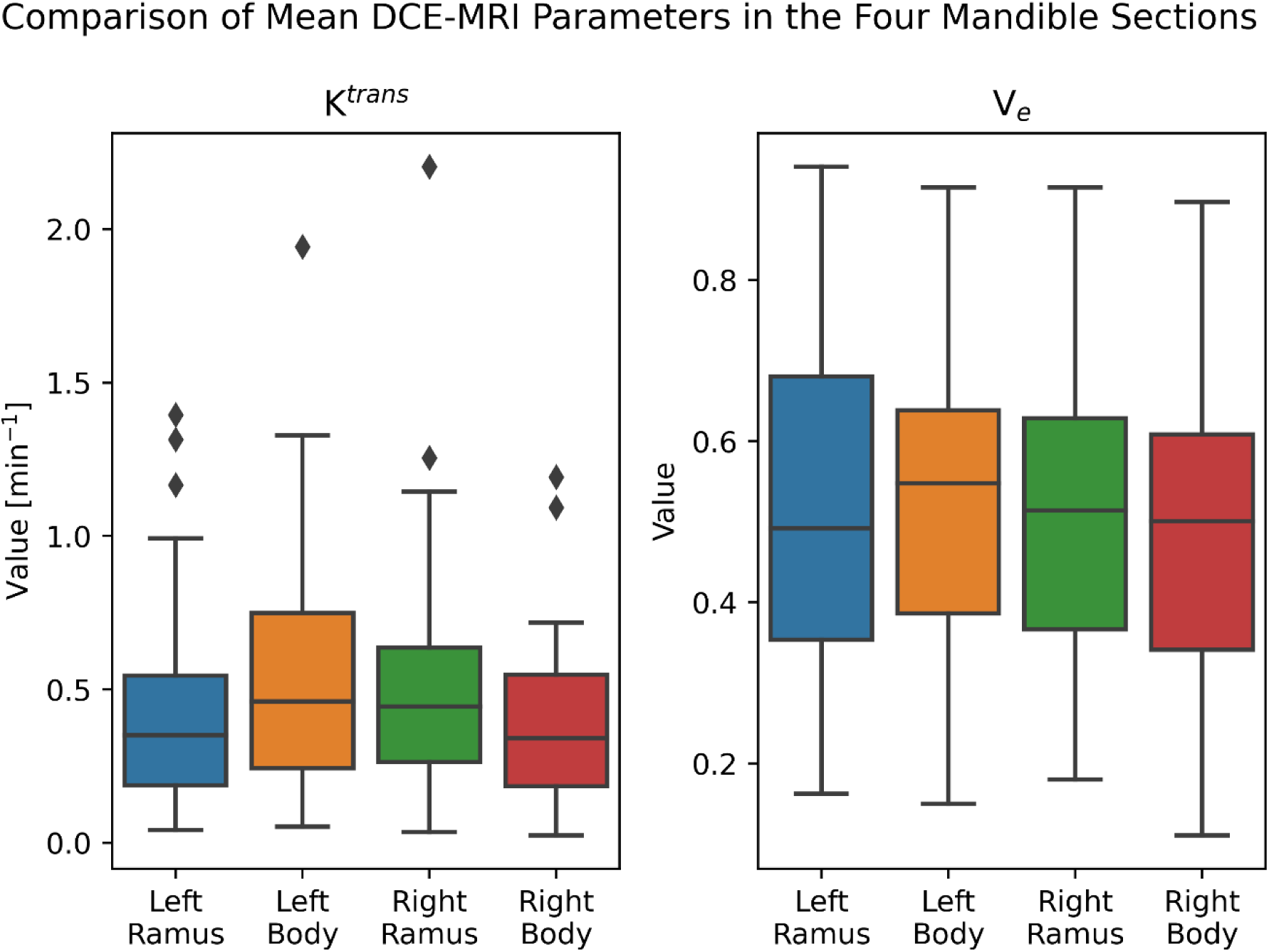
Comparison of DCE-MRI between mandible regions. The two Kruskal-Wallis tests comparing the mean values in each of the four mandible sections determined that there was no significant difference (*P* > 0.0125) in both K^trans^ and v_e_ in different regions of the mandible.

The differences in mandibular K^trans^ means between the high-dose and low-dose regions of the mandible were not significant as determined by the Wilcoxon-signed rank test (α = 0.0125, *W*=392, *Z=*2.0*, p*=0.044) (Figure 3). However, the differences in mandibular v_e_ means between the high-dose and low-dose regions were significant (*W*=214, *Z=*3.85, *p*=0.00013) (Figure 3).

**FIGURE 3.**
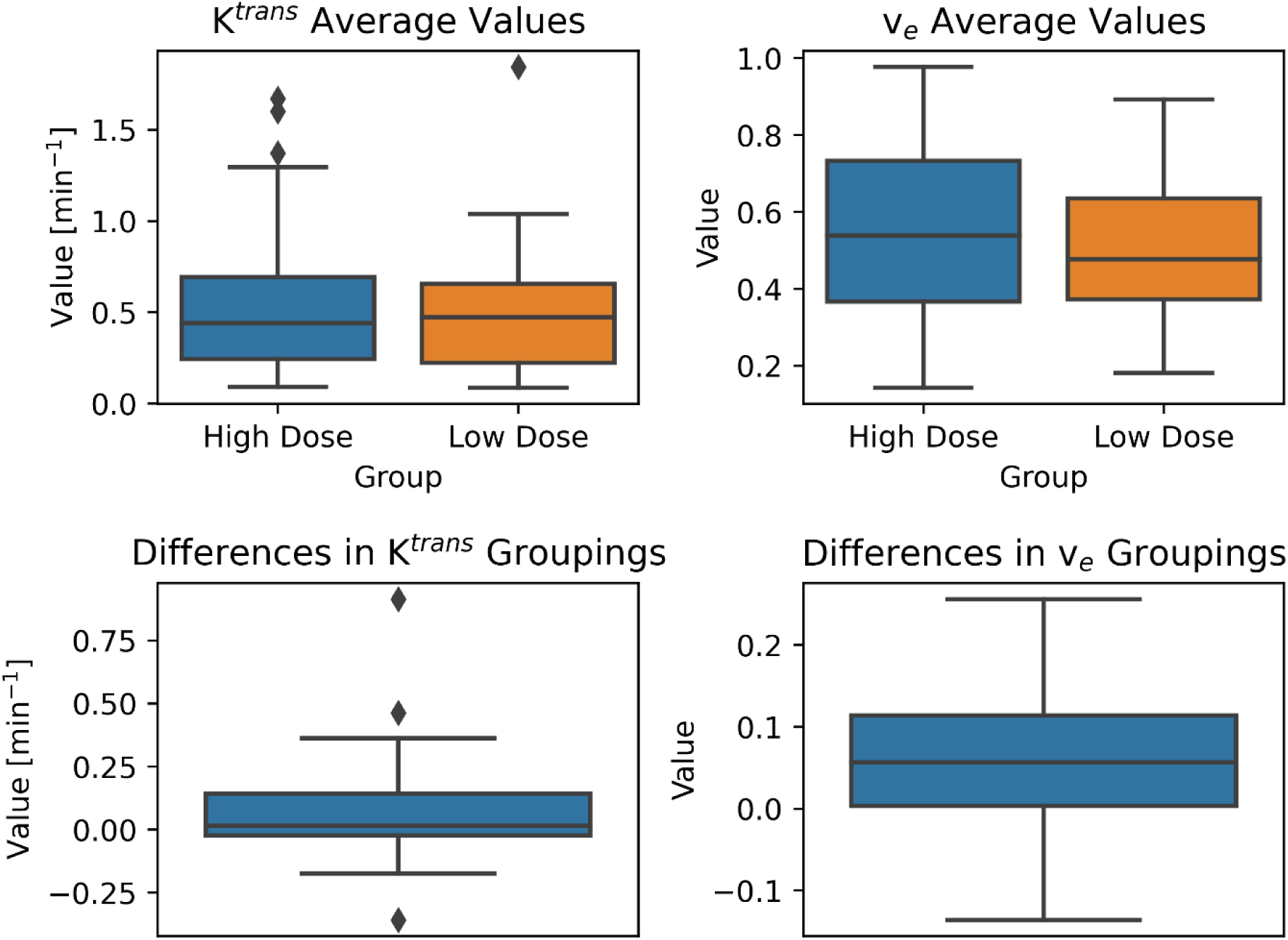
Summary of the DCE-MRI comparisons of the high- and low-dose mandible regions. The top two boxplots show the distribution of the calculated mean K^trans^ and v_e_ values in the images. The high dose boxes correspond to mean values of DCE-MRI parameters in mandible regions that received > 60 Gy. The low dose boxes correspond to mean values of DCE-MRI parameters in mandible regions that received ≤ 60 Gy. The bottom two boxplots show the DCE-MRI parameter differences between the high- and low-dose regions in the images. The differences are computed between the DCE-MRI parameter means of the high and low dose regions for the same patient. K^trans^ did not show a significant difference between high and low dose regions (*P* > 0.0125) whereas v_e_ did show a significant difference between high and low dose regions (*P* < 0.0125)

## DISCUSSION

The goal of this study was to determine if DCE-MRI can be used as an imaging biomarker for detecting mandibular physiological changes associated with high radiation dose from head and neck cancer radiation therapy.. F The results show that no inherent differences not attributed to high delivered radiation exist between the different mandible regions for either the K^trans^ or v_e_ parameters. Next, the high and low dose regions of the mandible were compared to determine if there are significant differences between DCE-MRI of the two regions. A statistically significant difference between the high and low dose regions of the mandible were identified for the v_e_ parameter, but not for the K^trans^ parameter. This parameter could be potentially used to identify radiation damage or osteoradionecrosis development within the mandible earlier compared to when symptoms presents clinically, allowing for earlier management of treatment-related symptoms.

The results of the high and low dose comparison were different between the two DCE-MRI variables studied: The K^trans^ parameter was not significantly different between the high and low dose mandible regions whereas the v_e_ parameter was significantly different between the high and low dose mandibular regions. [15]If the measured K^trans^ and v_e_ correspond to the physiological parameters specified by the Tofts model, the results indicate that the high dose regions have similar permeability to low dose regions but differing EES volumes.

There are several interpretations of the different results between the K^trans^ and v_e_ parameters. First, the radiation associated mandible changes in the EES may occur earlier than vascular permeability changes. Radiation therapy has shown to induce vascular changes such as inflammation and altered permeability in tissue[28][29][30][31][32]. It is possible that vascular permeability changes may be detectable much longer post-treatment rather than immediately following treatment such as with EES changes. These changes could be evaluated in future studies examining post-treatment DCE-MRI dynamics. Additionally, it is possible that tissue changes such as fibrosis, cell density, and other effects that result from radiation therapy could alter the mandible EES to a larger extent than mandible vascular permeability changes. A greater difference in v_e_ parameter means between high and low dose regions compared to K^trans^ parameter means may be easier to measure using DCE-MRI.

Several choices were made regarding the registration and resampling of the images. In this study, a rigid registration was used to register the T2w image and the CT instead of a deformable image registration. The overall purpose of the image registration was to align the DCE-MRI images to the treatment dose, specifically in the mandibular area. This approach was deemed appropriate due to minimal deformation in bone structures such as the mandible. Next, the treatment dose spacing was resampled to the spacing of the DCE-MRI instead of resampling the DCE-MRI spacing to the treatment dose spacing. This was completed because the dose was used only as a binary mask whereas the DCE-MRI values were used in the analysis.

This work has implicit limitations. First, there might be some uncertainty between the DCE-MRI and treatment plan registration. If there is a slight misregistration, the continuous nature of the dose maps should limit the impact on the results. Next, a single delineation of the mandible regions was completed for the mandible DCE-MRI comparisons. For this comparison, the consistency of the delineation of mandible regions is most important. Although the mandible region delineations were completed as accurately as possible, the results showing no inherent differences between mandible regions should remain if consistent, slight delineation deviations from actual mandible structures exist. Finally, a single dose (60 Gy) was used to distinguish between high and low dose regions. This dose was chosen due to it being on the lower end of the common HNC-RT prescription range of 60-70 Gy[33][34].

The work presented here complements other studies investigating the relationship between radiation dose and DCE-MRI in the mandible[11][23][24][25][26][27]. In a study conducted with a rabbit cohort, the change in K^trans^ and v_e_ of the mandible pre-radiation to post-radiation had no significant difference in the parameters between a control group that was not irradiated versus an experimental group that was irradiated[25]. However, there was a significant difference in DCE-MRI parameter changes between rabbits that received a mandible surgical procedure post-radiation compared to rabbits that received a mandible surgical procedure without prior radiation[25]. In comparison to this study, our work examined human post-treatment DCE-MRI rather than rabbit changes in DCE-MRI pre- and post-RT. Another study completed an analysis looking at the voxel-wise changes in DCE-MRI pre-RT and post-RT and found a significant difference in both the K^trans^ and v_e_ parameters[22]. Our study examined post-RT images rather than the change between pre-RT and post-RT and looked at differences between mandible regions rather than voxel-wise changes[22]. Finally, one study compared K^trans^ and v_e_ parameters of radiation therapy associated with osteoradionecrosis (ORN) affected regions of the mandible to control regions on the opposite side of the mandible[24]. It was found that there was a significant difference in K^trans^ and v_e_ in ORN affected volumes compared to the contralateral control volumes[24]. In addition, no correlation was found between the mean dose, min dose, max dose, and dose delivered to 95% of the ORN- ROIs between different subjects[24]. In comparison to that study, this study looked at DCE-MRI differences within each subject’s mandible between high and low dose regions and not DCE-MRI differences between ORN+ and ORN- affected regions[24]. Although not all regions that receive a large radiation dose will develop ORN, several studies have shown that high dose delivery to mandibular regions is a risk factor for ORN development[35][36][37].

Nonetheless, this effort supports the observed alteration of DCE parameters post-therapy as an indicator, and presents data supportive of DCE MRI as a response biomarker[38]. Future efforts are underway to formalize biomarker assessment as a monitoring biomarker of mandibular radiation injury, leading to consequential ORN in observational cohorts[39]. This work, by establishing a dichotomized (high/low) dose-response in a pilot cohort provides justification for further efforts at construction of imaging biomarker-informed normal tissue complication probability models which incorporate DCE MRI metrics.

## CONCLUSION

This work investigated whether a significant difference in post-RT DCE-MRI quantitative parameters exists between regions of the mandible receiving a high and low radiation dose. A significant difference between high and low dose regions of the mandible was observed for v_e_ whereas no significant difference was found for K^trans^. Determining whether a significant difference in these parameters may exist for regions at risk for developing ORN due to radiation damage may motivate development and validation of clinically relevant imaging-based biomarkers. Evaluating alterations in DCE-MRI parameters as a surrogate for radiation damage could identify patients at risk for ORN allowing for earlier treatment interventions for ORN and related toxicities associated with head and neck cancer radiation therapy.

## Data Availability

In accordance with NOT-OD-21-013, Final NIH Policy for Data Management and Sharing, anonymized/de-identified data that support the findings of this study are openly available in an NIH-supported generalist scientific data repository (figshare) at https://doi.org/10.6084/m9.figshare.29627075.v1 https://doi.org/10.6084/m9.figshare.29646260.v1 no later than the time of an associated publication.

https://doi.org/10.6084/m9.figshare.29646260.v1

## Data Sharing Statement

In accordance with NOT-OD-21-013, *Final NIH Policy for Data Management and Sharing*, anonymized/de-identified data that support the findings of this study are openly available in an NIH-supported generalist scientific data repository (figshare) at https://doi.org/10.6084/m9.figshare.29627075.v1 https://doi.org/10.6084/m9.figshare.29646260.v1 no later than the time of an associated publication.

## Public access policy compliance

In accordance with NOT-OD-25-049, *Supplemental Guidance to the 2024 NIH Public Access Policy: Government Use License and Rights*,: “This manuscript is the result of funding in whole or in part by the National Institutes of Health (NIH). It is subject to the NIH Public Access Policy. Through acceptance of this federal funding, NIH has been given a right to make this manuscript publicly available in PubMed Central upon the Official Date of Publication, as defined by NIH.”

## CRediT statement

In accordance with the Contributor Roles Taxonomy (CRediT, https://credit.niso.org/), the contributing authors have designated responsibilities and individual author attribution. The corresponding author(s) assume(s) responsibility for role assignment, and all contributors have been given the opportunity to review and confirm assigned roles. <u>Brandon Reber</u>: Methodology, Formal analysis, Writing-Original Draft, <u>Renjie He</u>: Software, Resources, <u>Moamen R Abdelaal</u>: Data Curation, <u>Abdallah Mohamed</u>: Data Curation, <u>Sam Mulder</u>: Writing-Review and Editing, <u>Laia Humbert Vidan</u>: Writing-Review and Editing, <u>Clifton Fuller</u>: Conceptualization, Resources, <u>Stephen</u> <u>Lai</u>: Conceptualization, Resources, Patient Trial Supervision, <u>Kristy Brock</u>: Conceptualization, Methodology, Supervision.

## Institutional Review Board Statement

Institutional-review board of the University of Texas MD Anderson Cancer Center, Houston, TX, USA gave ethical approval of this work under protocol PA16-0302. The protocol is available at clinicaltrials.gov (NCT03145077).

## Informed Consent Statement

Informed consent was collected from all patients, and all relevant informed consent forms are archived.

## REFERENCES

1. Sung, H.; Ferlay, J.; Siegel, R.L.; Laversanne, M.; Soerjomataram, I.; Jemal, A.; Bray, F. Global Cancer Statistics 2020: GLOBOCAN Estimates of Incidence and Mortality Worldwide for 36 Cancers in 185 Countries. CA Cancer J Clin 2021, *71*, 209–249, doi:10.3322/CAAC.21660.

2. Johnson, D.E.; Burtness, B.; Leemans, C.R.; Wai, V.; Lui, Y.; Bauman, J.E.; Grandis, J.R. Head and Neck Squamous Cell Carcinoma. 2021, doi:10.1038/s41572-020-00224-3.

3. Lydiatt, W.M.; Patel, S.G.; O’Sullivan, B.; Brandwein, M.S.; Ridge, J.A.; Migliacci, J.C.; Loomis, A.M.; Shah, J.P. Head and Neck Cancers—Major Changes in the American Joint Committee on Cancer Eighth Edition Cancer Staging Manual. CA Cancer J Clin 2017, 67, 122–137, doi:10.3322/CAAC.21389.

4. Parkin, D.M. Global Cancer Statistics in the Year 2000. Lancet Oncol 2001, 2, 533–543, doi:10.1016/S1470-2045(01)00486-7.

5. Chow, L.Q.M. Head and Neck Cancer. New England Journal of Medicine 2020, 382, 60–72, doi:10.1056/NEJMRA1715715.

6. Marur, S.; Forastiere, A.A. Head and Neck Cancer: Changing Epidemiology, Diagnosis, and Treatment. Mayo Clin Proc 2008, 83, 489–501, doi:10.4065/83.4.489.

7. Pfister, D.G.; Ang, K.K.; Brizel, D.M.; Burtness, B.A.; Busse, P.M.; Caudell, J.J.; Cmelak, A.J.; Colevas, A.D.; Dunphy, F.; Eisele, D.W.;, et al. Head and Neck Cancers, Version 2.2013. Journal of the National Comprehensive Cancer Network 2013, 11, 917–923, doi:10.6004/JNCCN.2013.0113.

8. Muzumder, S.; Srikantia, N.; Udayashankar, A.H.; Kainthaje, P.B.; John Sebastian, M.G.; Raj, J.M. Late Toxicities in Locally Advanced Head and Neck Squamous Cell Carcinoma Treated with Intensity Modulated Radiation Therapy. Radiat Oncol J 2021, 39, 184, doi:10.3857/ROJ.2020.00913.

9. Owosho, A.A.; Tsai, C.J.; Lee, R.S.; Freymiller, H.; Kadempour, A.; Varthis, S.; Sax, A.Z.; Rosen, E.B.; Yom, S.H.K.; Randazzo, J.;, et al. The Prevalence and Risk Factors Associated with Osteoradionecrosis of the Jaw in Oral and Oropharyngeal Cancer Patients Treated with Intensity-Modulated Radiation Therapy (IMRT): The Memorial Sloan Kettering Cancer Center Experience. Oral Oncol 2017, 64, 44–51, doi:10.1016/J.ORALONCOLOGY.2016.11.015.

10. Oh, H.K.; Chambers, M.S.; Martin, J.W.; Lim, H.J.; Park, H.J. Osteoradionecrosis of the Mandible: Treatment Outcomes and Factors Influencing the Progress of Osteoradionecrosis. J Oral Maxillofac Surg 2009, 67, 1378–1386, doi:10.1016/J.JOMS.2009.02.008.

11. Singh, A.; Huryn, J.M.; Kronstadt, K.L.; Yom, S.H.K.; Randazzo, J.R.; Estilo, C.L. Osteoradionecrosis of the Jaw: A Mini Review. Frontiers in Oral Health 2022, 3, 980786, doi:10.3389/FROH.2022.980786/BIBTEX.

12. Camolesi, G.C.V.; Ortega, K.L.; Medina, J.B.; Campos, L.; Pouso, A.I.L.; Vila, P.G.; Sayáns, M.P. Therapeutic Alternatives in the Management of Osteoradionecrosis of the Jaws. Systematic Review. Med Oral Patol Oral Cir Bucal 2020, 26, e195, doi:10.4317/MEDORAL.24132.

13. Petralia, G.; Summers, P.E.; Agostini, A.; Ambrosini, R.; Cianci, R.; Cristel, G.; Calistri, L.; Colagrande, S. Dynamic Contrast-Enhanced MRI in Oncology: How We Do It. Radiologia Medica 2020, 125, 1288–1300, doi:10.1007/S11547-020-01220-Z/FIGURES/7.

14. Gaddikeri, S.; Gaddikeri, R.S.; Tailor, T.; Anzai, Y. Dynamic Contrast-Enhanced MR Imaging in Head and Neck Cancer: Techniques and Clinical Applications. AJNR Am J Neuroradiol 2016, 37, 588, doi:10.3174/AJNR.A4458.

15. Tofts, P.S.; Kermode, A.G. Measurement of the Blood-Brain Barrier Permeability and Leakage Space Using Dynamic MR Imaging. 1. Fundamental Concepts. Magn Reson Med 1991, 17, 357–367, doi:10.1002/MRM.1910170208.

16. Kim, J.H.; Jenrow, K.A.; Brown, S.L. Mechanisms of Radiation-Induced Normal Tissue Toxicity and Implications for Future Clinical Trials. Radiat Oncol J 2014, 32, 103, doi:10.3857/ROJ.2014.32.3.103.

17. Nepon, H.; Safran, T.; Reece, E.M.; Murphy, A.M.; Vorstenbosch, J.; Davison, P.G. Healing, Inflammation, and Fibrosis: Radiation-Induced Tissue Damage: Clinical Consequences and Current Treatment Options. Semin Plast Surg 2021, 35, 181, doi:10.1055/S-0041-1731464.

18. Bülbül, H.M.; Bülbül, O.; Sarıoğlu, S.; Özdoğan, Ö.; Doğan, E.; Karabay, N. Relationships Between DCE-MRI, DWI, and 18F-FDG PET/CT Parameters with Tumor Grade and Stage in Patients with Head and Neck Squamous Cell Carcinoma. Mol Imaging Radionucl Ther 2021, 30, 177, doi:10.4274/MIRT.GALENOS.2021.25633.

19. Surov, A.; Meyer, H.J.; Gawlitza, M.; Höhn, A.K.; Boehm, A.; Kahn, T.; Stumpp, P. Correlations Between DCE MRI and Histopathological Parameters in Head and Neck Squamous Cell Carcinoma. Transl Oncol 2017, 10, 17–21, doi:10.1016/J.TRANON.2016.10.001.

20. Ota, Y.; Liao, E.; Kurokawa, R.; Syed, F.; Baba, A.; Kurokawa, M.; Moritani, T.; Srinivasan, A. Diffusion-Weighted and Dynamic Contrast-Enhanced MRI to Assess Radiation Therapy Response for Head and Neck Paragangliomas. Journal of Neuroimaging 2021, 31, 1035– 1043, doi:10.1111/JON.12875.

21. Bouten, R.M.; Young, E.F.; Selwyn, R.; Iacono, D.; W. Bradley Rittase; Day, R.M. Effects of Radiation on Endothelial Barrier and Vascular Integrity. *Tissue Barriers in Disease*, Injury and Regeneration 2021, 43–94, doi:10.1016/B978-0-12-818561-2.00007-2.

22. Dynamic Contrast-Enhanced MRI Detects Acute Radiotherapy-Induced Alterations in Mandibular Microvasculature: Prospective Assessment of Imaging Biomarkers of Normal Tissue Injury. Scientific Reports 2016 6:1 2016, 6, 1–11, doi:10.1038/srep29864.

23. Van Cann, E.M.; Rijpkema, M.; Heerschap, A.; van der Bilt, A.; Koole, R.; Stoelinga, P.J.W. Quantitative Dynamic Contrast-Enhanced MRI for the Assessment of Mandibular Invasion by Squamous Cell Carcinoma. Oral Oncol 2008, 44, 1147–1154, doi:10.1016/J.ORALONCOLOGY.2008.02.009.

24. Mohamed, A.S.R.; He, R.; Ding, Y.; Wang, J.; Fahim, J.; Elgohari, B.; Elhalawani, H.; Kim, A.D.; Ahmed, H.; Garcia, J.A.;, et al. Quantitative Dynamic Contrast-Enhanced MRI Identifies Radiation-Induced Vascular Damage in Patients With Advanced Osteoradionecrosis: Results of a Prospective Study. International Journal of Radiation Oncology*Biology*Physics 2020, 108, 1319–1328, doi:10.1016/J.IJROBP.2020.07.029.

25. Piotrowski, S.L.; Wilson, L.; Maldonado, K.L.; Tailor, R.; Hill, L.R.; Bankson, J.A.; Lai, S.; Kasper, F.K.; Young, S. Effect of Radiation on DCE-MRI Pharmacokinetic Parameters in a Rabbit Model of Compromised Maxillofacial Wound Healing: A Pilot Study. Journal of Oral and Maxillofacial Surgery 2020, 78, 1034.e1–1034.e10, doi:10.1016/J.JOMS.2020.02.001.

26. He, R.; Ding, Y.; Mohamed, A.S.R.; Ng, S.P.; Ger, R.B.; Elhalawani, H.; Elgohari, B.A.; Young, K.H.; Hutcheson, K.A.; Fuller, C.;, et al. Simultaneously Spatial and Temporal Higher-Order Total Variations for Noise Suppression and Motion Reduction in DCE and IVIM. Proc SPIE Int Soc Opt Eng 2020, 11313, 91, doi:10.1117/12.2549625.

27. Studer, G.; Bredell, M.; Studer, S.; Huber, G.; Glanzmann, C. Risk Profile for Osteoradionecrosis of the Mandible in the IMRT Era. Strahlentherapie Und Onkologie 2015, 192, 32, doi:10.1007/S00066-015-0875-6.

28. Bouten, R.M.; Young, E.F.; Selwyn, R.; Iacono, D.; W. Bradley Rittase; Day, R.M. Effects of Radiation on Endothelial Barrier and Vascular Integrity. *Tissue Barriers in Disease*, Injury and Regeneration 2021, 43–94, doi:10.1016/B978-0-12-818561-2.00007-2.

29. Wijerathne, H.; Langston, J.C.; Yang, Q.; Sun, S.; Miyamoto, C.; Kilpatrick, L.E.; Kiani, M.F. Mechanisms of Radiation-Induced Endothelium Damage: Emerging Models and Technologies. Radiother Oncol 2021, 158, 21, doi:10.1016/J.RADONC.2021.02.007.

30. Dekker, H.; Bravenboer, N.; van Dijk, D.; Bloemena, E.; Rietveld, D.H.F.; ten Bruggenkate, C.M.; Schulten, E.A.J.M. The Irradiated Human Mandible: A Quantitative Study on Bone Vascularity. Oral Oncol 2018, 87, 126–130, doi:10.1016/J.ORALONCOLOGY.2018.10.030.

31. Deshpande, S.S.; Donneys, A.; Farberg, A.S.; Tchanque-Fossuo, C.N.; Felice, P.A.; Buchman, S.R. Quantification and Characterization of Radiation-Induced Changes to Mandibular Vascularity Using Micro-Computed Tomography. Ann Plast Surg 2014, 72, 100, doi:10.1097/SAP.0B013E318255A57D.

32. Sønstevold, T.; Johannessen, C.A.; Stuhr, L. A Rat Model of Radiation Injury in the Mandibular Area. Radiat Oncol 2015, 10, doi:10.1186/S13014-015-0432-6.

33. Pignon, J.P.; Bourhis, J.; Domenge, C.; Designé, L.; Hill, C.; Adelstein, D.J.; Bachaud, J.M.; Bezwoda, W.R.; Buffoli, A.; Browman, G.P.;, et al. Chemotherapy Added to Locoregional Treatment for Head and Neck Squamous-Cell Carcinoma: Three Meta-Analyses of Updated Individual Data. The Lancet 2000, 355, 949–955, doi:10.1016/S0140-6736(00)90011-4.

34. Alfouzan, A.F. Radiation Therapy in Head and Neck Cancer. Saudi Med J 2021, 42, 247, doi:10.15537/SMJ.2021.42.3.20210660.

35. Aarup-Kristensen, S.; Hansen, C.R.; Forner, L.; Brink, C.; Eriksen, J.G.; Johansen, J. Osteoradionecrosis of the Mandible after Radiotherapy for Head and Neck Cancer: Risk Factors and Dose-Volume Correlations. 10.1080/0284186X.2019.1643037 2019, *58*, 1373–1377, doi:10.1080/0284186X.2019.1643037.

36. Pereira, I.F.; Firmino, R.T.; Meira, H.C.; Egito Vasconcelos, B.C. Do; Vladimir-Reimar-Augusto-De Souza, N.; Santos, V.R. Osteoradionecrosis Prevalence and Associated Factors: A Ten Years Retrospective Study. Med Oral Patol Oral Cir Bucal 2018, 23, e633, doi:10.4317/MEDORAL.22310.

37. Nabil, S.; Samman, N. Risk Factors for Osteoradionecrosis after Head and Neck Radiation: A Systematic Review. Oral Surg Oral Med Oral Pathol Oral Radiol 2012, 113, 54–69, doi:10.1016/J.TRIPLEO.2011.07.042.

38. Response Biomarker - BEST (Biomarkers, EndpointS, and Other Tools) Resource – NCBI Bookshelf Available online: https://www.ncbi.nlm.nih.gov/books/NBK402286/ (accessed on 27 June 2025).

39. Monitoring Biomarker - BEST (Biomarkers, EndpointS, and Other Tools) Resource – NCBI Bookshelf Available online: https://www.ncbi.nlm.nih.gov/books/NBK402282/ (accessed on 27 June 2025).

